# Epigenetic clock acceleration is linked to age-at-onset of idiopathic and *LRRK2* Parkinson’s disease

**DOI:** 10.1101/2022.02.04.22270486

**Authors:** Xuelin Tang, Paulina Gonzalez-Latapi, Connie Marras, Naomi P. Visanji, Wanli Yang, Christine Sato, Anthony E. Lang, Ekaterina Rogaeva, Ming Zhang

**Affiliations:** The First Rehabilitation Hospital of Shanghai, Department of Medical Genetics, School of Medicine, Tongji University, 200090, Shanghai, China; Edmond J. Safra Program in Parkinson’s disease and Morton and Gloria Shulman Movement Disorders Clinic, Toronto Western Hospital, Toronto, Canada; Ken and Ruth Davee Department of Neurology, Northwestern University, Feinberg School of Medicine, Chicago, Illinois, USA; Division of Neurology, University of Toronto, Toronto, Ontario, Canada; Tanz Centre for Research in Neurodegenerative Diseases, University of Toronto, 60 Leonard Ave., Toronto, ON, Canada, M5T 2S8; Department of Laboratory Medicine and Pathobiology, University of Toronto, Toronto, Ontario, Canada; Krembil Brain Institute, Toronto, Ontario, Canada; Clinical Center for Brain and Spinal Cord Research, Tongji University, Shanghai, China; Institute for Advanced Study, Tongji University, Shanghai, China

**Keywords:** *LRRK2*, mutation, epigenetics, DNA methylation, Parkinson’s disease

## Abstract

Parkinson’s disease is a clinically and genetically heterogeneous movement disorder with highly variable age-at-onset. DNA methylation (DNAm) age is an epigenetic clock that could reflect biological aging. Studies of DNAm-age acceleration (difference between DNAm-age and chronological age) are pertinent to neurodegenerative diseases (e.g., Parkinson’s disease), for which aging is the strongest risk-factor. We assessed DNAm-age in idiopathic Parkinson’s disease (n=96) and a longitudinal *LRRK2* cohort at four time-points over a 3-year period (n=220), including manifesting (n=91) and non-manifesting (n=129) G2019S-carriers. A highly variable age-at-onset was observed in both the idiopathic cohort (26-77 years) and manifesting G2019S-carriers (39-79 years). Increased DNAm-age acceleration was significantly associated with younger onset in idiopathic and *LRRK2*-related Parkinson’s disease, suggesting that every 5-year increase in DNAm-age acceleration is linked to about 6-year earlier onset. At an individual level, DNAm-age acceleration remained steady over a 3-year period for most G2019S-carriers, indicating that it might serve as a stable biomarker of biological aging. Future studies should evaluate the stability of DNAm-age acceleration over longer time-periods, especially for phenoconverters from non-manifesting to manifesting subjects. In conclusion, DNAm-age acceleration is linked to disease onset, and could be used in disease-modifying clinical trials of prodromal Parkinson’s disease.

## Introduction

Parkinson’s disease is a clinically heterogeneous neurodegenerative movement disorder characterized by tremor, rigidity, bradykinesia^1^ and highly variable age-at-onset ranging from juvenile to geriatric onset (up to 80 years apart)^2^. Parkinson’s disease is also genetically heterogeneous and the current catalog of genome-wide association studies (GWASs) include ∼90 independent loci associated with risk of idiopathic Parkinson’s disease.^3^ Additionally, approximately 10% of the patients have a familial form of the Parkinson’s disease. The most common causal mutation resides in *LRRK2* (p.G2019S; rs34637584) accounting for up to 2% of all Parkinson’s disease patients and being highly prevalent among patients of Ashkenazi Jewish origin (∼20%) and Arab-Berbers (∼40%). *LRRK2* mutations are linked to autosomal dominant Parkinson’s disease with incomplete age-dependent penetrance.^4^ A world-wide assessment revealed that the risk of Parkinson’s disease for G2019S-carriers was 28% at age 59, 51% at age 69, and 74% at age 79.^5^ Accordingly, G2019S-carriers have a variable age-at-onset, ranging between 50 and 80 years even within the same kindred.^6^

Even for familial Parkinson’s disease, our knowledge of possible disease modifiers is limited. Non-genetic modifiers, such as use of nonsteroidal anti-inflammatory drugs may be linked with reduced penetrance of Parkinson’s disease in *LRRK2*-carriers,^7,8^ however longitudinal observational and interventional studies are needed to confirm this association. In addition, an investigation of Arab-Berber G2019S-carriers suggested that the *DNM3* locus might be a genetic modifier of age-at-onset,^9^ though this finding has yet to be replicated in independent cohorts. Finally, a recent GWAS searched for genetic modifiers of disease penetrance or age-at-onset using the largest *LRRK2* cohort with 1879 participants, 96% of which were G2019S-carriers.^4^ Penetrance was associated with polygenic risk score and intronic variants in *CORO1C*, however modifiers of age-at-onset were not detected. Therefore, the goal of the present study was to search for age-at-onset modifiers of *LRRK2*-related and idiopathic Parkinson’s disease using an epigenetic approach.

Like genetic variations, epigenetic changes can alter gene expression, however epigenetic regulation is dynamic and can be influenced by many factors (e.g., environment, sex, and aging). Epigenetic mechanisms are known to affect cellular function,^10^ and clinical variability of Parkinson’s disease could be linked to epigenetic factors. One of the main epigenetic mechanisms is DNA methylation (DNAm), which plays a crucial role in normal development, genome stability, cell proliferation and aging. DNA methyltransferase adds a methyl group to cytosine residues mostly at CpG-sites, and DNAm levels at some CpGs are age-related. The cumulative evaluation of selected age-related CpGs can be used to estimate epigenetic clocks assessing DNAm-age, which we recently reviewed for utility in studying neurodegenerative diseases, including Parkinson’s disease.^11^ Intriguingly, several reports support the link between epigenetic clocks and biological age, the pace of which varies among individuals and may predict distinct aspects of aging at different life stages.^11^ The multi-tissue Horvath epigenetic clock^12^ is well-suited to study neurodegenerative diseases because it provides similar age predictions for most human tissues, including blood and brain. It consists of ∼350 CpGs mapped to 344 genes mainly involved in cell death, survival, and development.

The epigenetic clock is particularly relevant to neurodegenerative diseases, for which aging is the strongest risk-factor.^11^ In combination with genetic determinants of disease (e.g., G2019S in *LRRK2*), the epigenetic clock might help explain why some individuals stay healthy longer than others. Hence, study of the epigenetic clock has significant clinical relevance, including potentially a more accurate prediction of age-at-onset for preventive clinical trials. DNAm-age acceleration (difference between DNAm-age and chronological age) is associated with several neurodegenerative diseases,^11^ including age-at-onset of amyotrophic lateral sclerosis^13,14^. For Parkinson’s disease, one case-control study suggested the association of disease risk with DNAm-age acceleration.^15^ However, there are no reports assessing its link with Parkinson’s disease age-at-onset, except for a study of a single family with the causal p.A53E mutation in *SNCA*, which showed that an earlier onset was accompanied by greater DNAm-age acceleration.^16^ Notably, it is unknown if acceleration of DNAm-age is a cause or a consequence of neurodegeneration and how it changes with disease progression; therefore, longitudinal and prodromal cohorts are needed to evaluate DNAm-age over time. For instance, a longitudinal DNAm study of discordant identical twins revealed that the twin with amyotrophic lateral sclerosis had aged faster than the asymptomatic twin.^17^

Here we assessed DNAm-age in 96 patients with idiopathic Parkinson’s disease, and 220 G2019S-carriers at up to four time-points, including 129 non-manifesting carriers, which represent an opportune high-risk population to study prodromal Parkinson’s disease.

## Materials and methods

### Participants

Informed consent was obtained from all study participants. The idiopathic Parkinson’s disease cohort consisted of 96 patients (69% males) diagnosed at Toronto Western Hospital using the UK PD Society Brain Bank Clinical Diagnostic Criteria.^18^ All these patients were free from the G2019S *LRRK2* mutation based on the genotypes obtained as reported previously.^6^ Mean age-at-onset was 56 years (range 26-77) with a median of 57 years (interquartile range 47-65). This cohort was investigated in accordance with University Health Network Research Ethics Board approved protocol (UHN-REB 08-0615-AE). Data for the *LRRK2* cohort was obtained from the Parkinson’s Progression Marker Initiative (PPMI) at baseline, as well as 12-, 24-, and 36-month follow-up. The aims and methodology of PPMI have been published.^19^ All our study participants were enrolled at the initial stage of PPMI and included 91 manifesting and 129 non-manifesting G2019S-carriers (Table 1). Among 220 *LRRK2* subjects, 175 individuals were without known family history of Parkinson’s disease and 45 individuals were from 19 families with a history of Parkinson’s disease. Manifesting G2019S-carriers have a disease duration up to 7 years and may have been treated with dopaminergic drugs at baseline. Clinical data included the Movement Disorder Society-Unified Parkinson Disease Rating Scale (MDS-UPDRS)^20^ Part I (Non-Motor Experiences of Daily Living), Part II (Motor Experiences of Daily Living) and Part III (Motor Examination); as well as Hoehn and Yahr stage (measurement of Parkinson’s disease motor and functional disability), and the Montreal Cognitive Assessment (MoCA) total scores. DaTScan striatal binding ratio (SBR) data at baseline was also included in the analysis. SBR was calculated from count densities obtained from bilateral caudate and putamen as target regions and the occipital cortex as a reference tissue [(target region/reference region) – 1].

**Table 1.** Sample characteristics of the *LRRK2* G2019S-carriers.

|  | Baseline |  | 3-year follow-up |  |
| --- | --- | --- | --- | --- |
|  | Manifesting | Non-manifesting | Manifesting | Non-manifesting |
| Number of cases | 91 | 129 | 86 | 120 |
| Sex (n males, %) | 43 (47.3%) | 58 (45.0%) | 40 (46.5%) | 50 (41.7%) |
| White (n) | 88 | 128 | 83 | 121 |
| Hispanic or Latino (n) | 22 | 110 | 21 | 9 |
| Age-at-onset (years) |  |  |  |  |
| Mean (range) | 58 (36-75) | NA | 58 (36-84) | NA |
| Median (IQR) | 59 (51-64) | NA | 59 (51-64) | NA |
| Age at sample collection (years) |  |  |  |  |
| Mean (range) | 63 (39-79) | 62 (47-82) | 66 (42-84) | 65 (52-85) |
| Median (IQR) | 64 (58-69) | 62 (56-66) | 66 (60-72) | 65 (59-68) |
| MDS-UPDRS (mean) |  |  |  |  |
| MDS-UPDRS Part I | 8.14 | 4.45 | 7.26 | 3.43 |
| MDS-UPDRS Part II | 7.03 | 0.96 | 7.85 | 1.08 |
| MDS-UPDRS Part III | 21.02 | 2.92 | 22.62 | 3.86 |
| Hoehn and Yahr (n) |  |  |  |  |
| Stage 0 | 1 | 123 | 1 | 78 |
| Stage 1 | 22 | 5 | 14 | 10 |
| Stage 2 | 62 | 0 | 52 | 5 |
| Stage 3 | 6 | 0 | 11 | 0 |
| Stage 4 | 0 | 0 | 0 | 0 |
| MoCA (mean) | NA | NA | 26.27 | 27.22 |
| DaTscan striatal binding ratio (mean) |  |  |  |  |
| left caudate | 1.90 | NA | NA | NA |
| right caudate | 1.88 | NA | NA | NA |
| left putamen | 0.76 | NA | NA | NA |
| right putamen | 0.78 | NA | NA | NA |
| IQR = Interquartile Range; NA = not applicable. |  |  |  |  |

### DNAm analyses

For each study participant, DNAm data of about 850,000 CpG-sites were obtained from the Infinium MethylationEPIC array (Illumina) generated using bisulfite converted blood DNA. DNAm data for the *LRRK2* cohort (n=220) were available from PPMI, while the idiopathic Parkinson’s disease cohort (n=96) was processed at the Microarray Facility at the Centre for Applied Genomics (Toronto, Canada). The raw DNAm data were normalized using the Quantile preprocess method^21^ and analyzed using the minfi package in R-project^22^. DNAm levels at age-related CpGs were analyzed using the Horvath’s DNAm-age calculator tool (https://dnamage.genetics.ucla.edu/) to estimate DNAm-age based on an elastic net regression model.^12^ DNAm-age acceleration was calculated as DNAm-age minus chronological age at sample collection.

### Statistics

To analyze the association between DNAm-age acceleration and age-at-onset, we used a cox proportional hazard regression model (R survival and survminer packages)^23^ adjusted for sex, relatedness, the difference between age at sample collection and age-at-onset (interval), as well as blood cell counts (CD8T, CD4T, B-cell, and granulocyte) estimated based on the advanced analysis mode of the DNAm-age calculator. The adjustment for blood cell counts was implemented based on reported differences in imputed blood cell counts between Parkinson’s disease patients and controls.^15^ To adjust for relatedness, we created an indicator number for each family and used the R frailty function.^24^ In Kaplan-Meier curve analysis, we classified participants in each *LRRK2* and idiopathic cohort into three subgroups: normal aging (DNAm-age acceleration between −3 and 3 years), slow aging (DNAm-age acceleration <-3 years) and fast aging (DNAm-age acceleration >3 years), according to a previous study.^12^ We also used multivariate linear regression to assess the association between DNAm-age acceleration and age-at-onset (adjusted for sex and interval), as previously reported.^14^ R project 4.1.0 was used for the statistical analysis. A p-value <0.05 was considered statistically significant.

## Data availability

The data related to the current work are available upon reasonable request.

## Results

### DNAm-age acceleration is stable for most individuals within a 3-year period

We observed that DNAm-age is older than chronological age (DNAm-age acceleration above zero) in most patients with idiopathic Parkinson’s disease (75%) and G2019S-carriers at baseline (78%) with a similar pattern in manifesting and non-manifesting individuals (80% and 76%, respectively) (supplementary Fig. 1). Since longitudinal changes in DNAm-age are largely unknown, we analyzed DNAm-age acceleration at four time-points (from baseline to 3-year follow-up), which were available for 82 manifesting and 115 non-manifesting G2019S-carriers. Notably, DNAm-age acceleration in G2019S-carriers was similar between the manifesting and non-manifesting subgroups at baseline or 3-year follow-up (supplementary Fig. 2). At the individual level, DNAm-age acceleration remained steady over a 3-year period for most manifesting or non-manifesting G2019S-carriers (Fig. 1A, 1B). Indeed, 90% of manifesting and 92% of non-manifesting individuals had a small (<3 years) standard deviation of DNAm-age acceleration at the four time-points (only one manifesting and two non-manifesting individuals had a standard deviation of >4 years).

**Fig 1.**
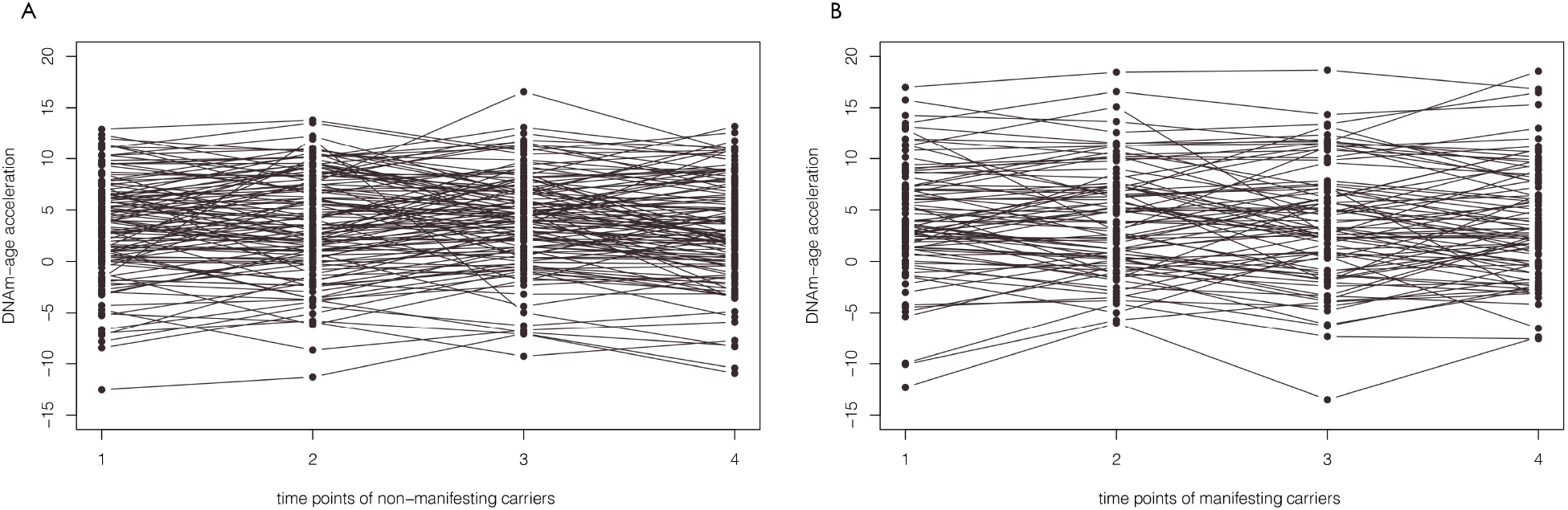
Line charts of DNAm-age acceleration at four time-points for each available G2019S-carrier: **A**. non-manifesting carriers (n=115), and **B**. manifesting carriers (n=82).

### DNAm-age acceleration is associated with age-at-onset of Parkinson’s disease

A highly variable age-at-onset was observed in either idiopathic Parkinson’s disease cohort (26-77 years; n=96) or manifesting G2019S-carriers (39-79 years; n=91) (Table 1). In the entire *LRRK2* cohort, cox proportional hazard regression analysis revealed a significant association of DNAm-age acceleration with age-at-onset at baseline (adjusted p=1.68E-6, HR=2.29, 95%CI:1.62-3.20, n=220) and the 3-year follow-up (adjusted p=1.65E-6, HR=2.56, 95%CI:1.74-3.75, n=206) (Fig. 2A, 2B; supplementary Table 1). These results suggest that faster aging group has an increased hazard for an earlier onset of 129% at baseline and 156% at 3-year follow-up. This association was also significant in manifesting G2019S-carriers (adjusted p=1.70E-10 HR=3.83, 95%CI:2.53-5.78) and idiopathic Parkinson’s disease cohort (adjusted p=0.0145, HR=1.53, 95%CI:1.09-2.16), with an increased hazard of 283% and 53%, respectively (Fig. 3A, 3B; supplementary Table 1). Specifically, in the fast *vs*. slow aging group, the median onset was 13 years earlier for idiopathic patients (59 *vs*. 72 years) and 14 years earlier for manifesting G2019S-carriers (59 *vs*. 73 years). For the entire *LRRK2* cohort, the median onset was 12 years earlier in the fast *vs*. slow aging group at baseline (65 *vs*. 77 years) or 3-year follow-up (69 *vs*. 81 years).

**Fig 2.**
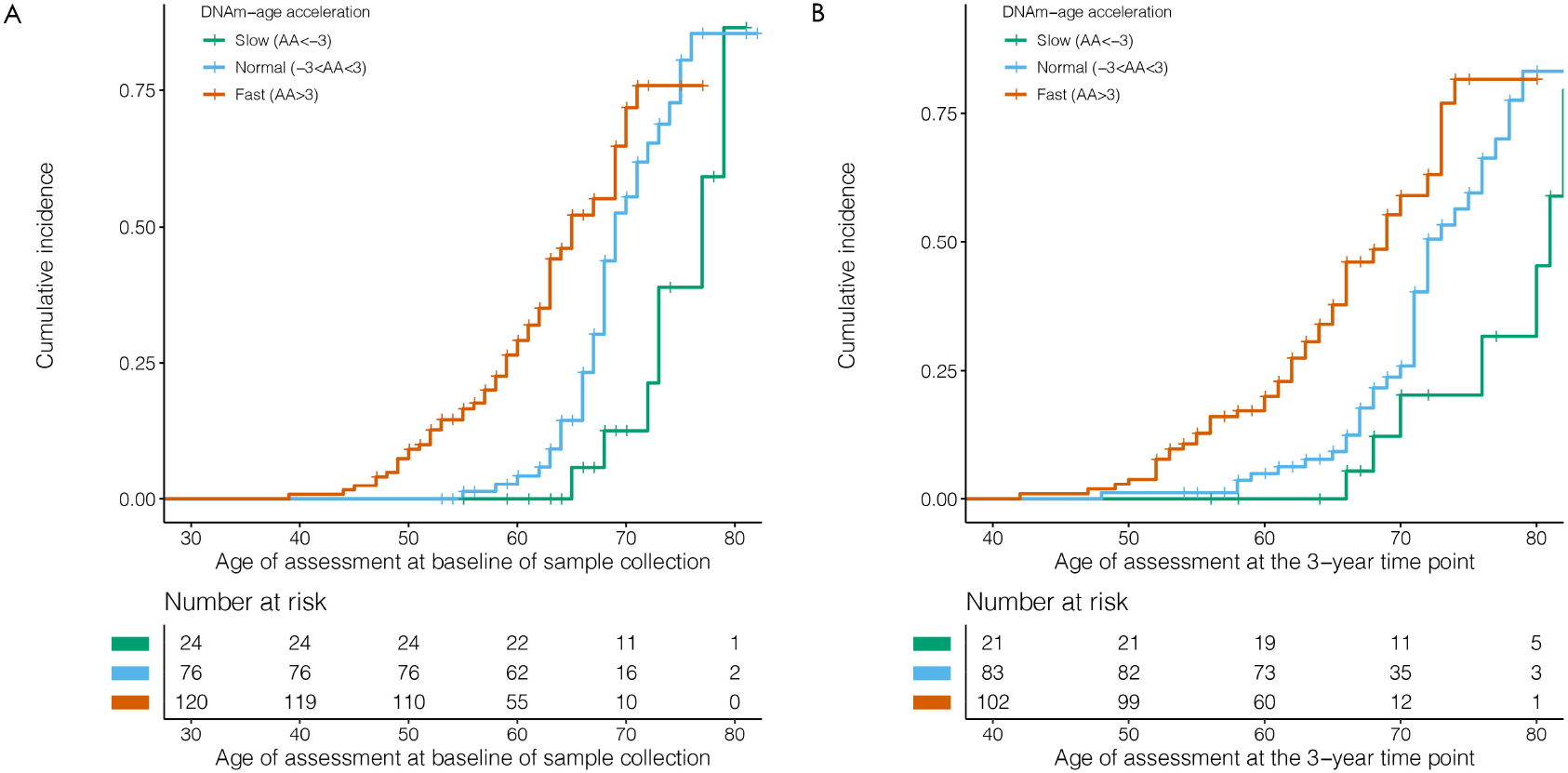
Kaplan-Meier curves for the *LRRK2* cohort. The effect of DNAm-age acceleration (AA<-3, - 3<AA<3, AA>3) on age-at-onset of Parkinson’s Disease: **A**. at baseline sample collection (n=220); p=1.68E-6, HR=2.29(95%CI:1.62-3.20), and **B**. 3-year follow-up (n=206); p=1.65E-6, HR=2.56(95%CI:1.74-3.75). Analyses were adjusted for sex, relatedness, and blood cell (CD8T, CD4T, B cell, and Gran cells).

**Fig 3.**
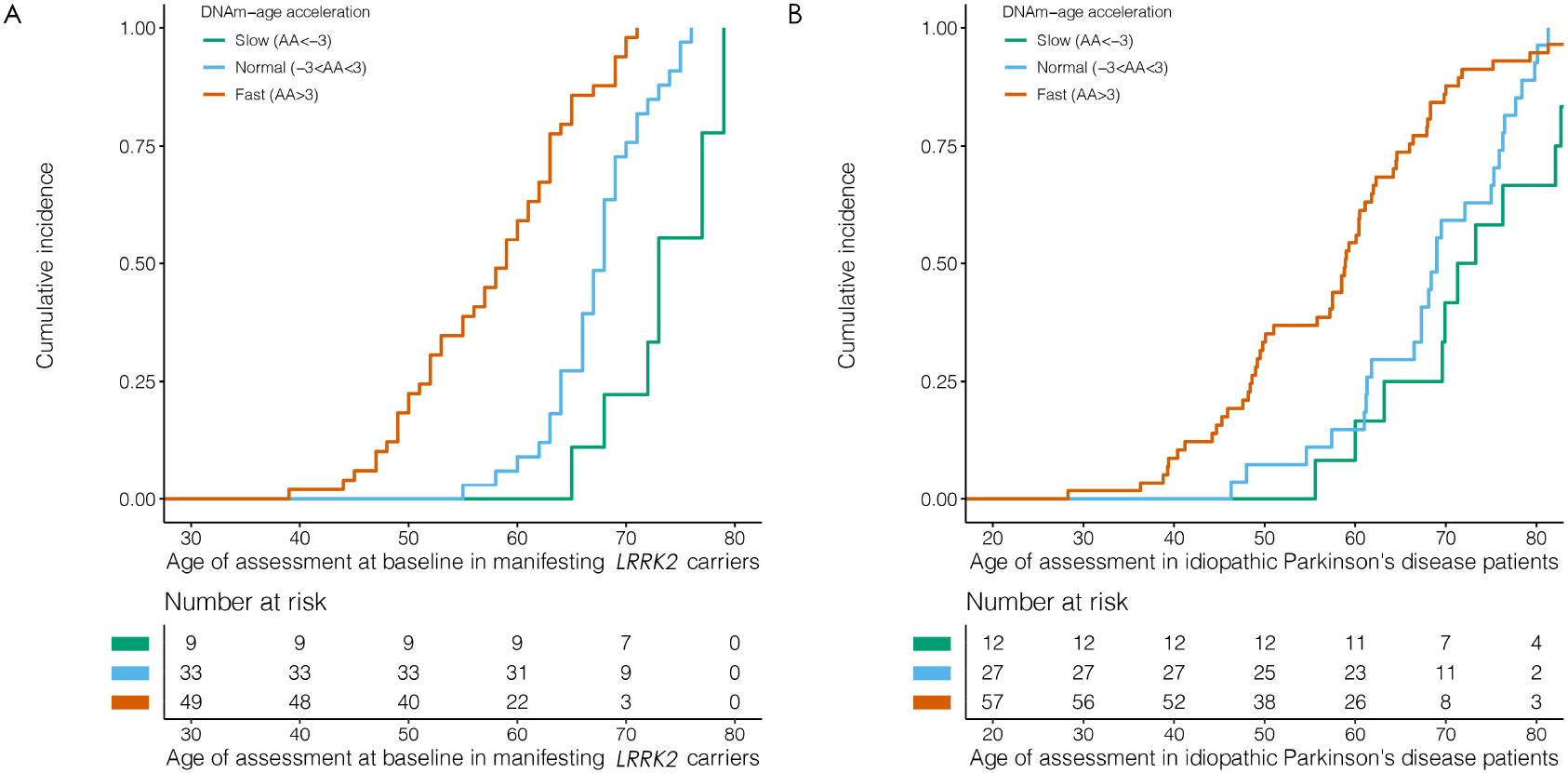
Kaplan-Meier curves showing the effect of DNAm-age acceleration (AA<-3, - 3<AA<3, AA>3) on age-at-onset of Parkinson’s Disease: **A**. at baseline sample collection in 91 manifesting G2019S-carriers; p=1.70E-10, HR=3.83(95%CI:2.53-5.78), and **B**. 96 idiopathic Parkinson’s disease patients; p=0.0145, HR=1.53(95%CI:1.09-2.16). Analyses were adjusted for sex, interval, and blood cell (CD8T, CD4T, B cell, and Gran cells).

The link between DNAm-age acceleration and age-at-onset was supported by multivariate linear regression analyses (adjusted for sex and interval) in manifesting G2019S-carriers at baseline (p=2.25E-15, B=-1.15, R^2^ =0.51, n=91) or 3-year follow-up (p=1.89E-14, B=-1.17, R^2^=0.52, n=86); as well as in the idiopathic Parkinson’s disease cohort (p=5.39E-9, B=-1.19, R^2^=0.30, n=96) (supplementary Fig. 3 A-C). These data suggest that every 5-year increase in DNAm-age acceleration is linked to 6-year earlier onset in G2019S-carriers or idiopathic patients. Of note, one of the non-manifesting G2019S-carriers (#54215) was diagnosed with Parkinson’s disease by the 3-year follow-up (supplementary Table 2). DNAm-age acceleration of this phenoconverter increased by 5.5 years from baseline (−6.7 years) to 2-year follow-up (−1.2 year), supporting its link with onset at early stage of the Parkinson’s disease (supplementary Fig. 4).

### DNAm-age acceleration is not linked with Parkinson’s disease severity, cognitive ability or dopamine transporter binding

In manifesting G2019S-carriers, we did not detect a significant association between DNAm-age acceleration and disease severity based on motor signs (MDS-UPDRS Part III), non-motor and motor symptoms (MDS-UPDRS Part I and II) at either baseline or 3-year follow-up (supplementary Fig. 5). Similarly, no association was observed with Hoehn and Yahr stage (supplementary Fig. 6) or MoCA score in G2019S-carriers at 3-year follow-up (supplementary Fig.7). In addition, we observed no significant association between DNAm-age acceleration and SBR data at baseline for different brain regions (p>0.05 for left caudate, right caudate, left putamen), except for the right putamen (p=0.046, R^2^=0.025, B=-0.016) (supplementary Fig. 8), indicating that increased DNAm-age acceleration was associated with a lower level of dopamine binding in that region.

## Discussion

Our study has revealed a significant association between increased DNAm-age acceleration and younger onset in both idiopathic and *LRRK2*-associated Parkinson’s disease, suggesting that every 5-year increase in DNAm-age acceleration is linked to about 6-year earlier onset. We did not detect an association with indicators of Parkinson’s disease severity, except for a suggestive link of increased DNAm-age acceleration with a lower level of dopamine binding in the right putamen, which needs to be validated in independent studies.

Our investigation of the *LRRK2* cohort included a unique longitudinal design, which allowed for the evaluation of DNAm-age at four time-points. For most manifesting and non-manifesting G2019S-carriers, the dynamics of DNAm-age acceleration was stable within a 3-year period, supporting that it might serve as a steady biomarker of biological aging linked to age-at-onset of Parkinson’s disease. However, measurement of DNAm-age over longer time-periods are necessary to establish the extent of this stability. Notably, one of the non-manifesting G2019S-carriers had phenoconverted to Parkinson’s disease by the 3-year follow-up, which was accompanied by 5.5-year increased DNAm-age acceleration within two years prior to disease onset. Future study of a larger sample set of phenoconverters should clarify the dynamics of DNAm-age acceleration around the time of onset of clinically manifest Parkinson’s disease. Notably, rapid-eye-movement sleep behavior disorder (RBD) is the strongest prodromal marker for α□synucleinopathies, including Parkinson’s disease.^25^ Therefore, the analysis of DNAm-age in RBD may contribute to the understanding of the factors linked to phenoconversion of RBD to Parkinson’s disease, which could vary between 2 and 12 years. A longitudinal study design is needed to evaluate DNAm-age during RBD progression; and investigate if acceleration of DNAm-age is the cause or consequence of neurodegeneration.

The underlying mechanisms behind the association of epigenetic clock with age-at-onset of Parkinson’s disease remain to be investigated. Single cell RNA-sequencing data of mouse/human brain tissues^26^ would allow for the assessment of whether disease-related genes^27,28^ or GWAS loci are enriched in specific cell types using cell-type enrichment tools (e.g., EWCE^29^). Similar investigation of age-related genes might also reveal specific brain cell types that are vulnerable to biological aging and explain variable age-at-onset in Parkinson’s disease and other neurodegenerative diseases. Modern technologies (e.g., whole-genome bisulfite sequencing) could detect more age-related CpGs in the human methylome for better tracking of biological aging. Future studies should also investigate aging mechanisms reflected by different DNAm clocks, each of which has its own strengths and limitations.^11^ Such studies might focus on the use of the most suitable DNAm clock in a disease-specific context and determine whether a combination of DNAm clocks is more useful to capture the complexity of aging and the clinical heterogeneity of age-related neurodegenerative diseases.

In conclusion, our study finds compelling data that the epigenetic clock may anticipate PD age-at-onset in both idiopathic and *LRRK2*-related Parkinson’s disease. DNAm-age acceleration might be a promising aging biomarker that is linked to Parkinson’s disease and other neurodegenerative disease. It is consistent in both blood and CNS tissues,^30^ and stable for most of our study participants during a 3-year period. DNAm clock(s) could be used in disease-modifying clinical trials of prodromal Parkinson’s disease. DNAm-age acceleration might be considered as an inclusion criterion when recruitment of individuals with a relatively high likelihood of manifesting Parkinson’s disease within the timeframe of the trial is desired.

## Data Availability

All data produced in the present study are available upon reasonable request to the authors.

## Acknowledgments

The authors would like to acknowledge the patients who participated in this research. Partial data used in the preparation of this article were obtained from the Parkinson’s Progression Markers Initiative (PPMI) database (www.ppmi-info.org/access-dataspecimens/download-data). For up-to-date information on the PPMI study, visit https://www.ppmi-info.org/. PPMI, a public-private partnership, is funded by the Michael J. Fox Foundation for Parkinson’s Research and funding partners, including: 4D Pharma, AbbVie Inc., AcureX Therapeutics, Allergan, Amathus Therapeutics, Aligning Science Across Parkinson’s (ASAP), Avid Radiopharmaceuticals, Bial Biotech, Biogen, BioLegend, Bristol Myers Squibb, Calico Life Sciences LLC, Celgene Corporation, DaCapo Brainscience, Denali Therapeutics, The Edmond J. Safra Foundation, Eli Lilly and Company, GE Healthcare, GlaxoSmithKline, Golub Capital, Handl Therapeutics, Insitro, Janssen Pharmaceuticals, Lundbeck, Merck & Co. Inc., Meso Scale Diagnostics, LLC, Neurocrine Biosciences, Pfizer Inc., Piramal Imaging, Prevail Therapeutics, F. Hoffmann□La Roche Ltd and its affiliated company Genentech Inc., Sanofi Genzyme, Servier, Takeda Pharmaceutical Company, Teva Neuroscience Inc., UCB, Vanqua Bio, Verily Life Sciences Inc., Yumanity Therapeutics Inc., Voyager Therapeutics.

## Reference

1. Kalia LV, Lang AE. Parkinson’s disease. Review. Lancet. Aug 29 2015;386(9996):896–912. doi:10.1016/S0140-6736(14)61393-3

2. Hill-Burns EM, Ross OA, Wissemann WT, et al. Identification of genetic modifiers of age-at-onset for familial Parkinson’s disease. Hum Mol Genet. Sep 1 2016;25(17):3849–3862. doi:10.1093/hmg/ddw206

3. Buniello A, MacArthur JAL, Cerezo M, et al. The NHGRI-EBI GWAS Catalog of published genome-wide association studies, targeted arrays and summary statistics 2019. Research Support, N.I.H., Extramural Research Support, Non-U.S. Gov’t. Nucleic Acids Res. Jan 8 2019;47(D1):D1005–D1012. doi:10.1093/nar/gky1120

4. Lai D, Alipanahi B, Fontanillas P, et al. Genomewide Association Studies of LRRK2 Modifiers of Parkinson’s Disease. Ann Neurol. Jul 2021;90(1):76–88. doi:10.1002/ana.26094

5. Healy DG, Falchi M, O’Sullivan SS, et al. Phenotype, genotype, and worldwide genetic penetrance of LRRK2-associated Parkinson’s disease: a case-control study. Multicenter Study Research Support, N.I.H., Extramural Research Support, Non-U.S. Gov’t Research Support, U.S. Gov’t, Non-P.H.S. Lancet Neurol. Jul 2008;7(7):583–90. doi:10.1016/S1474-4422(08)70117-0

6. Marras C, Klein C, Lang AE, et al. LRRK2 and Parkin mutations in a family with parkinsonism-Lack of genotype-phenotype correlation. Case Reports Research Support, Non-U.S. Gov’t. Neurobiol Aging. Apr 2010;31(4):721–2. doi:10.1016/j.neurobiolaging.2008.05.030

7. Yahalom G, Rigbi A, Israeli-Korn S, et al. Age at Onset of Parkinson’s Disease Among Ashkenazi Jewish Patients: Contribution of Environmental Factors, LRRK2 p.G2019S and GBA p.N370S Mutations. J Parkinsons Dis. 2020;10(3):1123–1132. doi:10.3233/JPD-191829

8. San Luciano M, Tanner CM, Meng C, et al. Nonsteroidal Anti-inflammatory Use and LRRK2 Parkinson’s Disease Penetrance. Mov Disord. Oct 2020;35(10):1755–1764. doi:10.1002/mds.28189

9. Trinh J, Gustavsson EK, Vilarino-Guell C, et al. DNM3 and genetic modifiers of age of onset in LRRK2 Gly2019Ser parkinsonism: a genome-wide linkage and association study. Lancet Neurol. Nov 2016;15(12):1248–1256. doi:10.1016/S1474-4422(16)30203-4

10. Kundaje A, Meuleman W, Ernst J, et al. Integrative analysis of 111 reference human epigenomes. Research Support, N.I.H., Extramural Research Support, Non-U.S. Gov’t Research Support, U.S. Gov’t, Non-P.H.S. Nature. Feb 19 2015;518(7539):317–30. doi:10.1038/nature14248

11. Bergsma T, Rogaeva E. DNA Methylation Clocks and Their Predictive Capacity for Aging Phenotypes and Healthspan. Review. Neurosci Insights. 2020;15:2633105520942221. doi:10.1177/2633105520942221

12. Horvath S. DNA methylation age of human tissues and cell types. Genome Biol. 2013;14(10):R115. doi:10.1186/gb-2013-14-10-r115

13. Zhang M, Tartaglia MC, Moreno D, et al. DNA methylation age-acceleration is associated with disease duration and age at onset in C9orf72 patients. Research Support, Non-U.S. Gov’t. Acta Neuropathol. Aug 2017;134(2):271–279. doi:10.1007/s00401-017-1713-y

14. Zhang M, McKeever PM, Xi Z, et al. DNA methylation age acceleration is associated with ALS age of onset and survival. Research Support, Non-U.S. Gov’t. Acta Neuropathol. May 2020;139(5):943–946. doi:10.1007/s00401-020-02131-z

15. Horvath S, Ritz BR. Increased epigenetic age and granulocyte counts in the blood of Parkinson’s disease patients. Research Support, N.I.H., Extramural. Aging (Albany NY). Dec 2015;7(12):1130–42. doi:10.18632/aging.100859

16. Picillo M, Lizarraga KJ, Friesen EL, et al. Parkinsonism due to A53E alpha-synuclein gene mutation: Clinical, genetic, epigenetic, and biochemical features. Research Support, Non-U.S. Gov’t. Mov Disord. Dec 2018;33(12):1950–1955. doi:10.1002/mds.27506

17. Zhang M, Xi Z, Ghani M, et al. Genetic and epigenetic study of ALS-discordant identical twins with double mutations in SOD1 and ARHGEF28. J Neurol Neurosurg Psychiatry. Nov 2016;87(11):1268–1270. doi:10.1136/jnnp-2016-313592

18. Hughes AJ, Daniel SE, Kilford L, Lees AJ. Accuracy of clinical diagnosis of idiopathic Parkinson’s disease: a clinico-pathological study of 100 cases. J Neurol Neurosurg Psychiatry. Mar 1992;55(3):181–4. doi:10.1136/jnnp.55.3.181

19. Parkinson Progression Marker I. The Parkinson Progression Marker Initiative (PPMI). Prog Neurobiol. Dec 2011;95(4):629–35. doi:10.1016/j.pneurobio.2011.09.005

20. Goetz CG, Tilley BC, Shaftman SR, et al. Movement Disorder Society-sponsored revision of the Unified Parkinson’s Disease Rating Scale (MDS-UPDRS): scale presentation and clinimetric testing results. Mov Disord. Nov 15 2008;23(15):2129–70. doi:10.1002/mds.22340

21. Touleimat N, Tost J. Complete pipeline for Infinium((R)) Human Methylation 450K BeadChip data processing using subset quantile normalization for accurate DNA methylation estimation. Epigenomics. Jun 2012;4(3):325–41. doi:10.2217/epi.12.21

22. Aryee MJ, Jaffe AE, Corrada-Bravo H, et al. Minfi: a flexible and comprehensive Bioconductor package for the analysis of Infinium DNA methylation microarrays. Bioinformatics. May 15 2014;30(10):1363–9. doi:10.1093/bioinformatics/btu049

23. Terry M Therneau PMG. Modeling survival data: extending the cox model.. Springer; 2000.

24. Zhang M, Ferrari R, Tartaglia MC, et al. A C6orf10/LOC101929163 locus is associated with age of onset in C9orf72 carriers. Brain. Oct 1 2018;141(10):2895–2907. doi:10.1093/brain/awy238

25. Postuma RB, Iranzo A, Hu M, et al. Risk and predictors of dementia and parkinsonism in idiopathic REM sleep behaviour disorder: a multicentre study. Brain. Mar 1 2019;142(3):744–759. doi:10.1093/brain/awz030

26. Ming GL, Song H. Seq-ing out cell types across the isocortex and hippocampal formation. Cell. Jun 10 2021;184(12):3083–3085. doi:10.1016/j.cell.2021.05.016

27. Skene NG, Bryois J, Bakken TE, et al. Genetic identification of brain cell types underlying schizophrenia. Nat Genet. Jun 2018;50(6):825–833. doi:10.1038/s41588-018-0129-5

28. Bryois J, Skene NG, Hansen TF, et al. Genetic identification of cell types underlying brain complex traits yields insights into the etiology of Parkinson’s disease. Nat Genet. May 2020;52(5):482–493. doi:10.1038/s41588-020-0610-9

29. Zeisel A, Muñoz-Manchado AB, Codeluppi S, et al. Brain structure. Cell types in the mouse cortex and hippocampus revealed by single-cell RNA-seq. Science (New York, NY). 2015;347(6226):1138–1142. doi:10.1126/science.aaa1934

30. Horvath S. DNA methylation age of human tissues and cell types. Genome Biol. 2013;14(10):R115. doi:10.1186/gb-2013-14-10-r115

