## Supplemental materials for "Epigenetic clock acceleration is linked to age-at-onset of idiopathic and *LRRK2* Parkinson’s disease"

**Supplementary Table 1. Summary of the cox proportional hazards analysis when considering DNAm-age acceleration as continuous or categorized (into three groups) variants.**

| | Continuous | Categorized [a] |
|---|---|---|
| *LRRK2* cohort at baseline* | | |
| p-value | 7.44E-10 | 1.68E-6 |
| HR | 1.16 | 2.29 |
| 95%CI | 1.11-1.21 | 1.62-3.20 |
| *LRRK2* cohort at 3-year follow-up* | | |
| p-value | 1.24E-10 | 1.65E-6 |
| HR | 1.19 | 2.56 |
| 95%CI | 1.13-1.26 | 1.74-3.75 |
| Manifesting G2019S-carriers at baseline△ | | |
| p-value | 2.19E-12 | 1.70E-10 |
| HR | 1.18 | 3.83 |
| 95%CI | 1.13-1.24 | 2.53-5.78 |
| Idiopathic Parkinson's Disease patients[#] | | |
| p-value | 3.76E-5 | 0.0145 |
| HR | 1.09 | 1.53 |
| 95%CI | 1.05-1.14 | 1.09-2.16 |

*Adjusted for sex, relatedness and blood cell count (CD8T, CD4T, B cells, and Gran cells).

△Adjusted for sex, relatedness, interval between age at onset and age at sample collection and blood cell count (CD8T, CD4T, B cells, and Gran cells).

[#] Adjusted for sex, interval between age at onset and age at sample collection and blood cell count (CD8T, CD4T, B cells, and Gran cells).

[a] we created different indicator number for each group (0 for slowing aging,1for normal aging and 2 for fast aging).

**Supplementary Table 2. Clinical characteristics of phenoconverter (G2019S-carriers #54215).**

| | Baseline | 1-year follow-up | 2-year follow-up | 3-year follow-up |
|---|---|---|---|---|
| MDS-UPDRS Part I | 6 | 10 | 10 | 10 |
| MDS-UPDRS Part II | 4 | 3 | 4 | 6 |
| MDS-UPDRS Part III | 4 | 15 | 25 | 30 |
| Hoehn & Yahr | Stage 0 | Stage 1 | Stage 0 | Stage 3 |
| MoCA | NA | 25 | 25 | 20 |
| DNAm-age acceleration values(years) | -6.65 | -3.92 | -1.22 | -2.07 |

NA = not applicable.

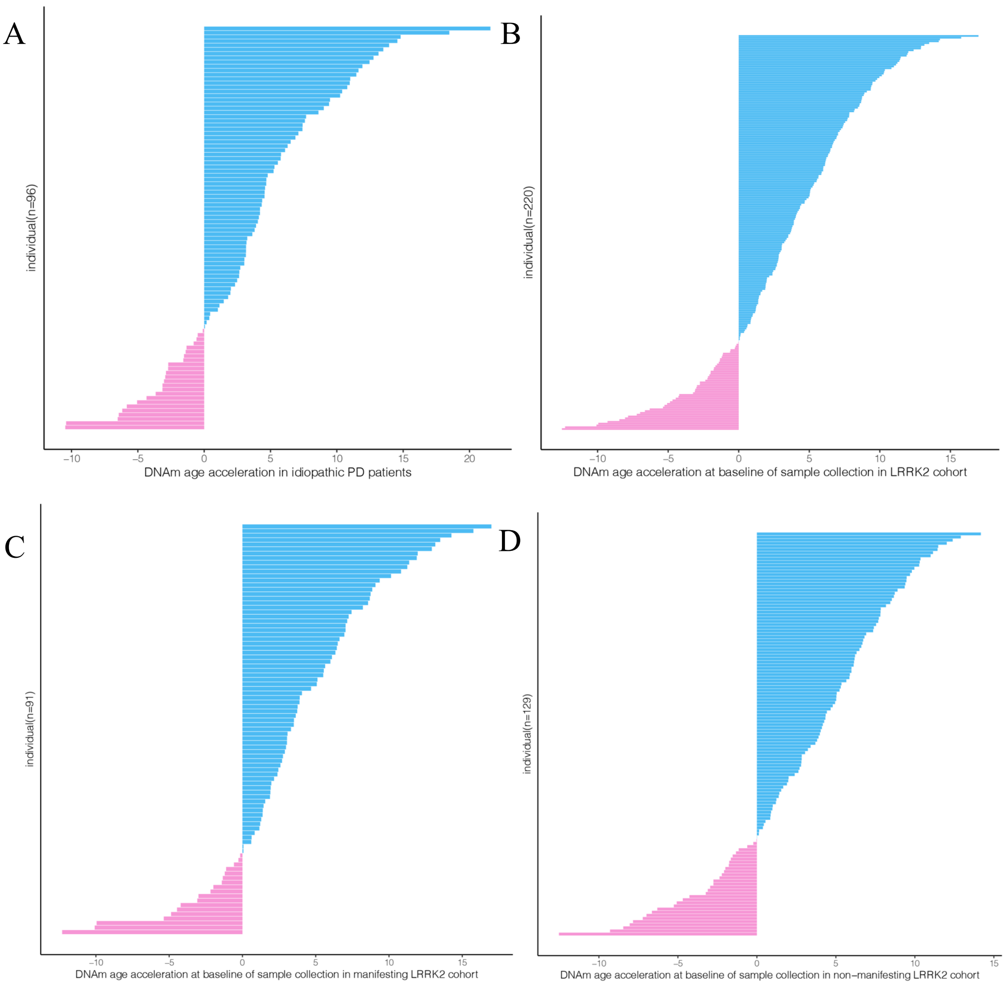


**Supplementary Fig 1.** The distribution of DNAm-age acceleration in 96 idiopathic PD patients **(A)**, 220 G2019S-carriers at baseline **(B)**, including 91 manifesting **(C)** and 129 non-manifesting **(D)** G2019S-carriers.

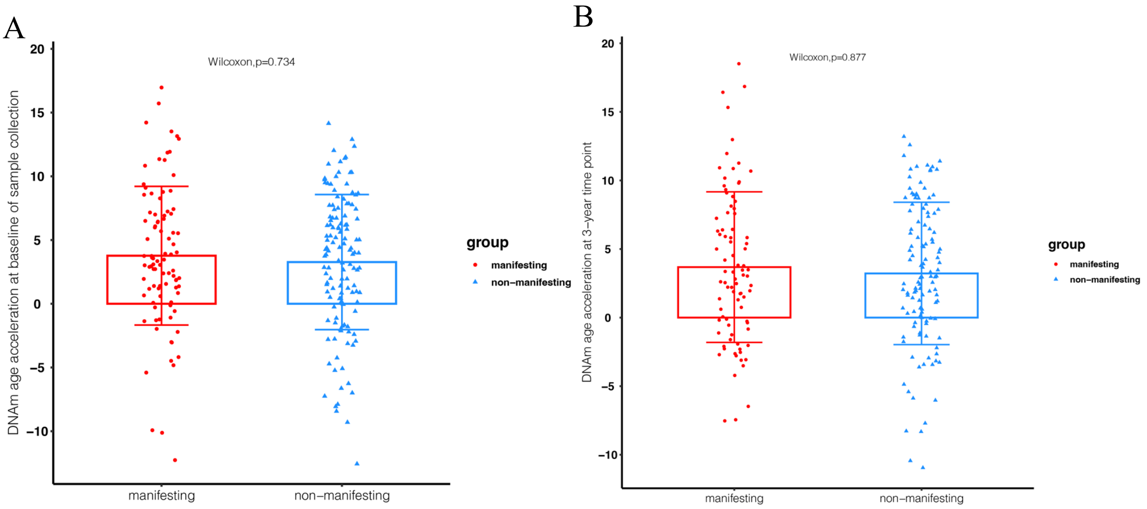


**Supplementary Fig 2.** Bar/scatter charts of DNAm-age acceleration in G2019S-carriers at baseline and the 3-year time-point of sample collection. There is no significant difference between DNAm-age acceleration for manifesting and non-manifesting G2019S-carriers at **A.** baseline (p=0.73, Wilcoxon test) or **B.** the 3-year time-point (p=0.88, Wilcoxon test).

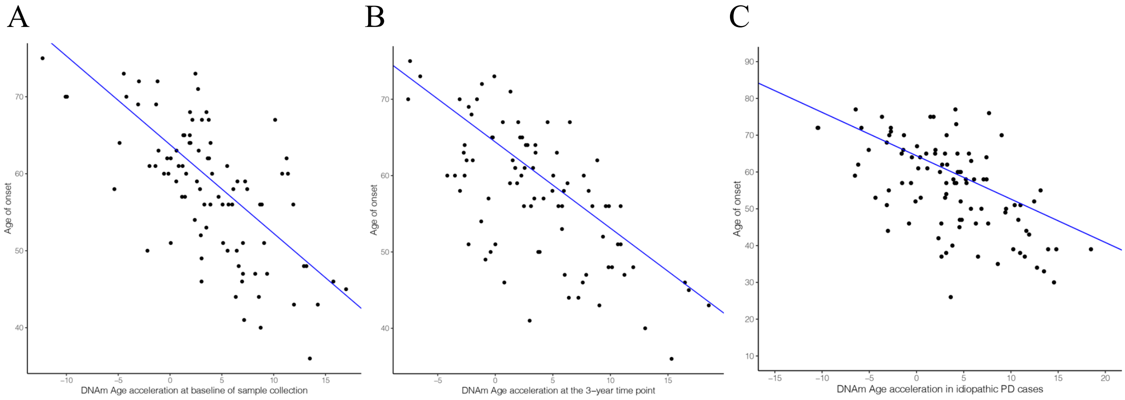


**Supplementary Fig 3.** Scatter plot of DNAm-age acceleration and age-at-onset in PD patients with or without an *LRRK2* mutation. The association between DNAm-age accelerationand age-at-onset in **A.** G2019S-carriers at baseline sample collection (p=2.25E-15, B=-1.15, $R^2$ =0.51, n=91); **B.** G2019S-carriers at 3-year follow-up (p=1.89E-14, B=-1.17, $R^2$ =0.52, n=86). **C.** idiopathic PD patients (p=5.39E-9, B=-1.19, $R^2$ =0.30, n=96). Linear regression analyses were adjusted for sex and interval.

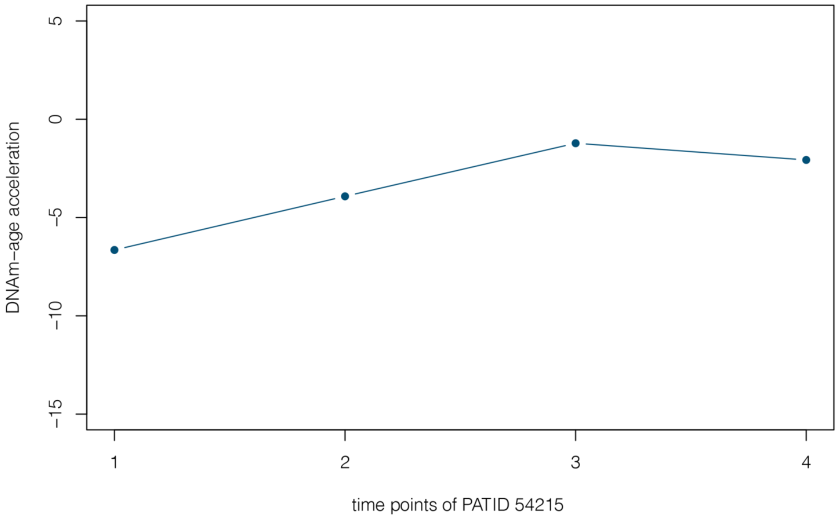


**Supplementary Fig. 4**. DNAm-age acceleration of the phenoconverter (patient #54215) at four time points, who was non-manifesting at up to 2-year follow-up (time-points 1-3), but was diagnosed with Parkinson’s Disease by 3-year follow-up (time-point 4).

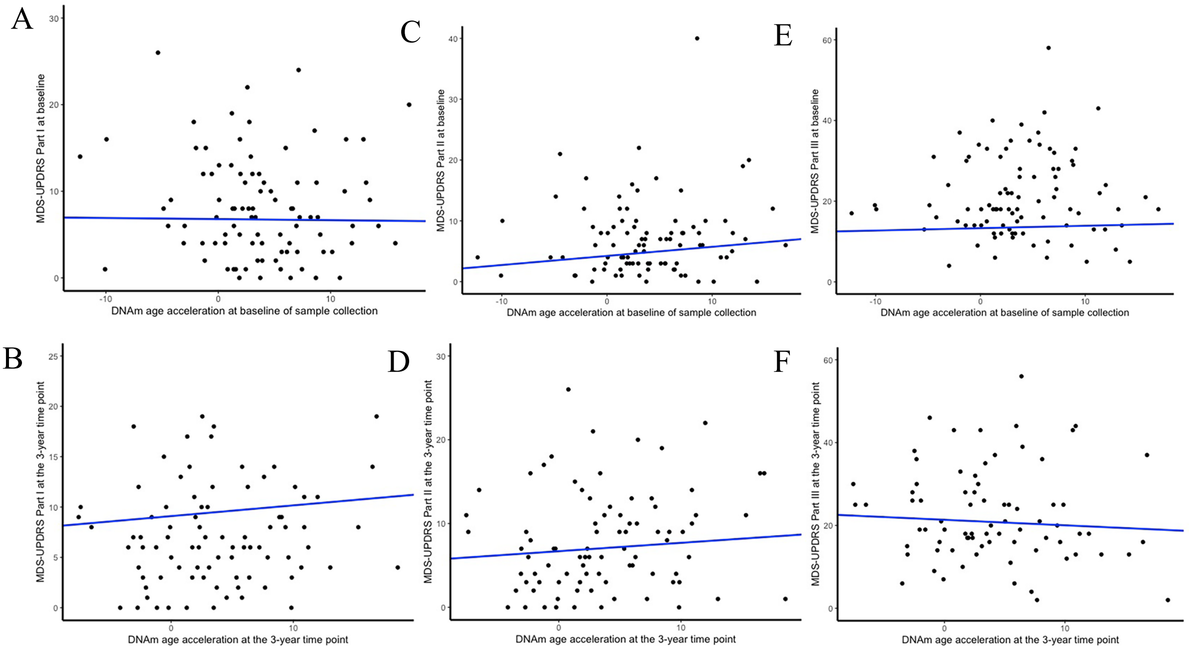


**Supplementary Fig 5.** Scatter plots of DNAm-age acceleration and UPDRS Part I-III at baseline (n=91) and the 3-year time point (n=85 for part I and part II, n=77 for part III) for *LRRK2* G2019S-carriers. DNAm-age acceleration is not significantly associated with UPDRS (p>0.05, adjusted for sex, family and interval).

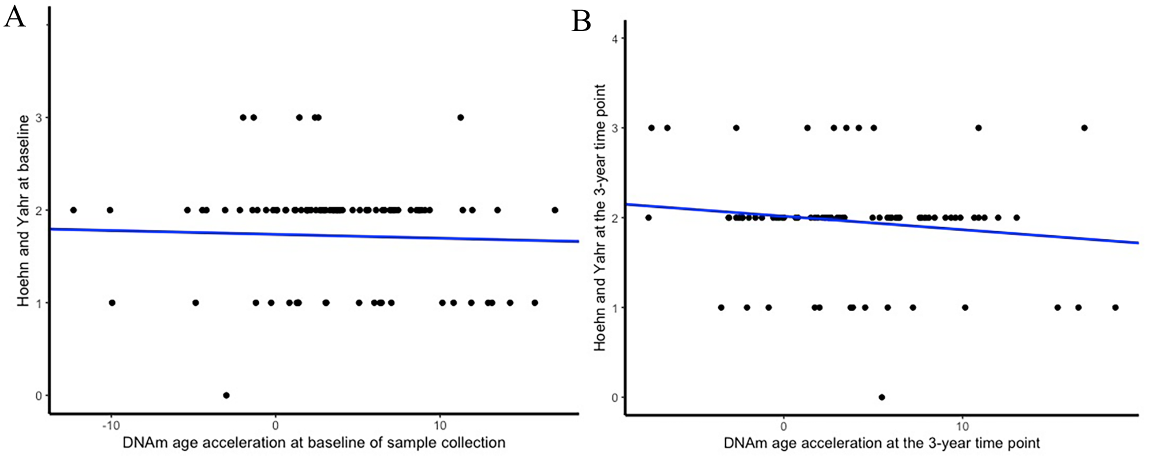


**Supplementary Fig 6.** Scatter plots of DNAm-age acceleration and the Hoehn and Yahr scale for *LRRK2* G2019S-carriers at **(A)** baseline (n=91, p>0.05) and **(B)** 3-year follow-up (n=77, p>0.05). P-values are adjusted for sex, relatedness and interval. The blue line represents the linear regression trend.

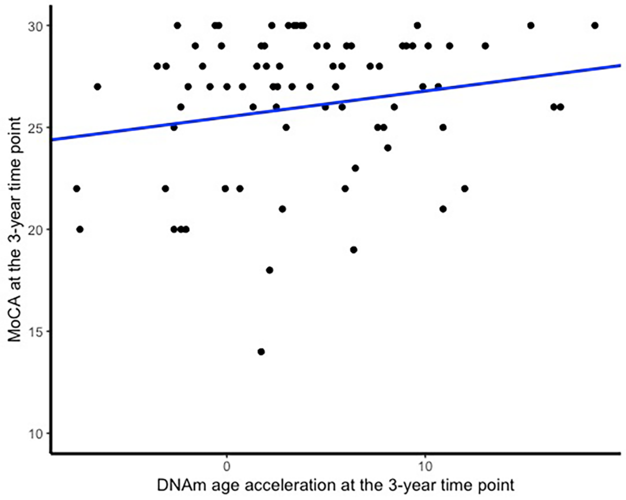


**Supplementary Fig 7.** Scatter plot of DNAm-age acceleration and MoCA score in G2019S-carriers at 3-year follow-up (p=0.079, $R^2$=0.019, B=0.13, adjusted for sex, n=78).

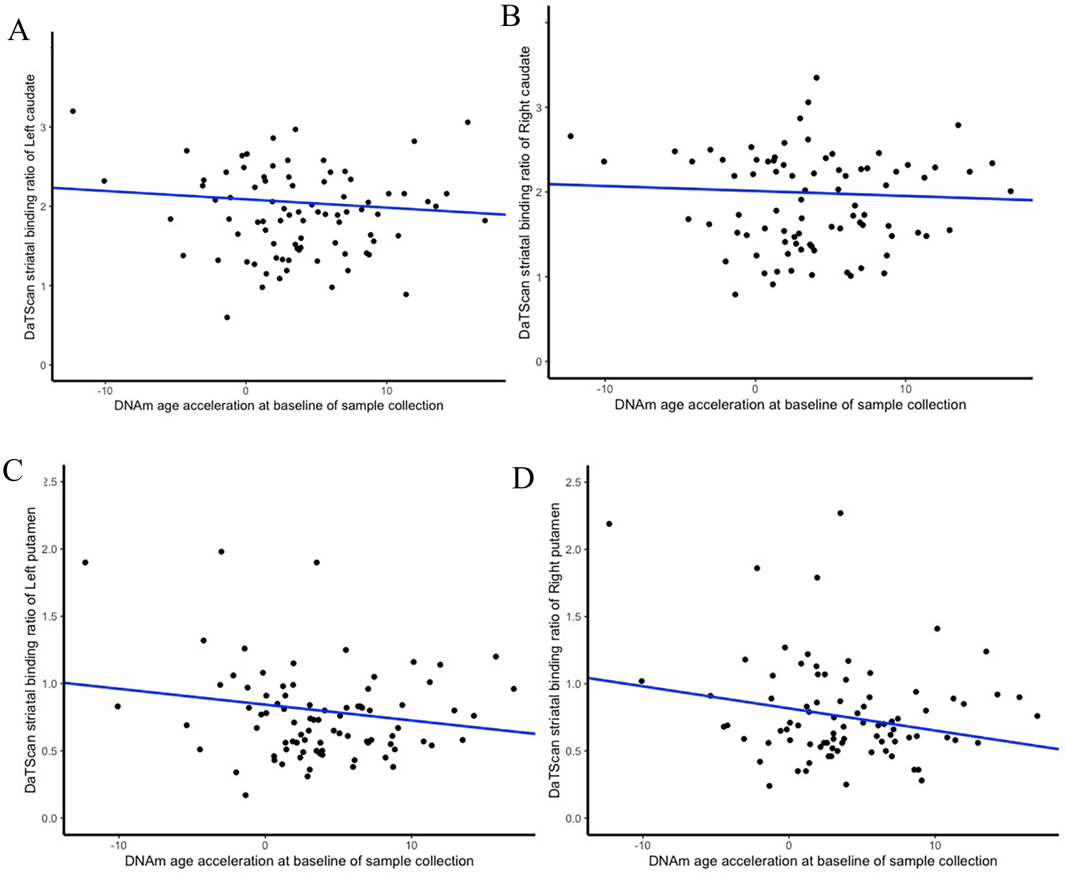


**Supplementary Fig 8.** Scatter plots of DNAm-age acceleration and DaTScan striatal binding ratio in the *LRRK2* G2019S-carriers (n=84) at baseline in **(A)** left caudate (p=0.35), **(B)** right caudate (p=0.63), **(C)** left putamen (p=0.081) and **(D)** right putamen (p=0.046, $R^2$=0.02451, B=-0.016). P-values were adjusted for sex, family and interval).
